# Variations in the Effect of Pulse Pressure on Cognition Based on Blood Pressure

**DOI:** 10.1101/2025.11.17.25340451

**Authors:** Mubarick Saeed, Charles F. Murchison, David S. Geldmacher, Katie M. Wheeler, Erik D. Roberson, Victor A. Del Bene

## Abstract

**Background:** An increase in pulse pressure, the difference between systolic and diastolic blood pressure, has emerged as a predictor of cognitive impairment in older adults. However, its predictive utility relative to blood pressure status and potential differential effects based on blood pressure status remain unclear.

**Methods:** This study used data from the ACTIVE trial (N = 2,802 adults aged 65+) to examine the association between pulse pressure and cognitive decline over 10 years. Linear mixed-effects models assessed the relationship between pulse pressure, blood pressure, and decline in global cognition, memory, reasoning, and processing speed.

**Results:** Higher pulse pressure was associated with faster global cognitive decline (*b*= −0.003, 95% CI [−0.004, −0.001]) after controlling for demographics, cognitive training status, and attrition. This effect was consistent for the memory and reasoning domains. The effects remained significant after adjusting for blood pressure status, with pulse pressure demonstrating complementary predictive utility to blood pressure. Specifically, the negative effect of pulse pressure on global cognitive decline was observed only among participants with elevated blood pressure (*b* = −0.005, 95% CI [−0.005, −0.002]), not in those with normal blood pressure. This pattern was consistent across the three cognitive domains.

**Conclusions:** Pulse pressure is associated with accelerated cognitive decline among older adults, but this relationship is moderated by blood pressure status. Pulse pressure monitoring could improve cognitive risk assessment in older adults with elevated blood pressure. This underscores the importance of managing blood pressure to mitigate the risk of cognitive decline associated with increased pulse pressure.

## Introduction

Hypertension is a known risk factor for cognitive impairment among older adults (Forte et al., 2020; Sierra, 2020) and pulse pressure, the difference between systolic and diastolic blood pressure, is an emerging predictor of cognitive impairment in older adults (Aimagambetova et al., 2024; Zhou et al., 2023). Pulse pressure reflects the force generated by the heart per beat and is a marker for arterial stiffness as indexed by pulse wave velocity (Muxfeldt et al., 2008). However, the predictive utility of pulse pressure relative to blood pressure status, whether normal or elevated, for cognitive decline is unclear. Additionally, while there is some evidence that the relationship between pulse pressure and cognitive decline might be influenced by blood pressure status, few studies have investigated this potential differential effect (McDade et al., 2016; Muela et al., 2018; Nation et al., 2016). Understanding this relationship is crucial for clinical practice, as pulse pressure may offer a more sensitive marker for cognitive risk stratification in older adults, especially among those with hypertension.

Higher pulse pressure among older adults is associated with both lower cognitive functioning and an elevated risk of cognitive impairment (Nation et al., 2016; Sha et al., 2018; Zhou et al., 2023) in addition to accelerated neurodegeneration, more rapid progression to dementia, and Alzheimer’s disease biomarkers (positive amyloid beta and phosphorylated tau) (McDade et al., 2016; Nation et al., 2015; Nation et al., 2016; Shi et al., 2020). However, Sha et al. (2018) found that the effect of pulse pressure on the rate of cognitive decline was non-significant after accounting for several covariates, including hypertension. Considering hypertension is also associated with cognitive decline, dementia, and brain atrophy (Alateeq et al., 2021; Tzourio et al., 1999), it is critical to understand whether pulse pressure has an independent effect on late-life cognition apart from blood pressure status.

There is mixed evidence as to whether pulse pressure provides additional predictive utility beyond blood pressure status. For instance, Blacher et al. (2000) found pulse pressure is superior to mean blood pressure in predicting cardiovascular complications. In contrast, other studies found pulse pressure is an inferior predictor of cardiovascular outcomes relative to systolic and diastolic blood pressure (Miura et al., 2001; Mosley et al., 2007). Considering the different cohorts of these studies, the conflicting findings suggest a differential effect of pulse pressure depending on blood pressure levels. Blacher et al. (2000) reported pulse pressure as a superior predictor in a cohort of hypertensive participants, while Miura et al. (2001) and Mosley et al. (2007) found pulse pressure to be an inferior predictor in a cohort of non-hypertensive participants. Prior studies have not explored this comparative predictive utility between pulse pressure and blood pressure status in cognitive outcomes.

Few studies have investigated the differential effect of pulse pressure on cognitive functioning based on blood pressure status. For instance, McDade et al. (2016) found the effect of pulse pressure on cognitive decline was greater among older adults with higher systolic blood pressure at baseline compared to those with lower systolic blood pressure. In contrast, Nation et al. (2016) found the interaction between pulse pressure and blood pressure status on cognitive decline was not significant. The limited number of studies and their conflicting findings warrant a more in-depth investigation of the research question in a larger cohort.

In the present study, we investigated the association between pulse pressure and cognitive decline. The first aim was to examine the effect of pulse pressure on cognitive decline among older adults. We then examined whether pulse pressure provides an additional predictive benefit of cognitive decline over that of blood pressure status. Finally, we evaluated whether there is a differential effect of pulse pressure on cognitive decline based on blood pressure status. We hypothesized that higher pulse pressure would be associated with faster cognitive decline, and this relationship would remain significant after controlling for blood pressure status. We further hypothesized that the effect of pulse pressure on cognitive decline would be stronger among participants with elevated blood pressure compared to those with normal blood pressure.

## Methods

The data that support the findings of this study are available from the corresponding author upon reasonable request. The data is also available at the National Archive of Computerized Data on Aging (https://www.icpsr.umich.edu/web/NACDA/studies/38821).

### Dataset

This secondary data analysis used data from the ACTIVE (Advanced Cognitive Training for Independent and Vital Elderly) study, a randomized, controlled clinical trial to promote independence among older adults through cognitive training (Jobe et al., 2001; Willis et al., 1998). The sample included 2802 older adults (aged 65+) enrolled from 6 sites across the United States, including the University of Alabama at Birmingham, Hebrew Rehabilitation Center for the Aged, Indiana University, Johns Hopkins University, Pennsylvania State University, and the University of Florida/Wayne State University. The ACTIVE study recruited healthy older adults with no substantial cognitive impairment at baseline. The present study excluded 15 ACTIVE participants due to self-reported Alzheimer’s disease at baseline. Baseline assessment included demographics, blood pressure readings, health history, and cognitive functioning. Follow-up assessments occurred at approximately the first, second, third, fifth, and tenth-year post-baseline assessment, covering blood pressure readings, health history, and cognitive functioning.

### Measures

#### Pulse Pressure and Blood Pressure Status

Participants underwent two consecutive blood pressure readings at each time point. The mean of the two blood pressure readings determined the systolic and diastolic blood pressure for each time point. Pulse pressure for each time point was calculated by the difference between systolic and diastolic blood pressure.

Participants were stratified into two groups by blood pressure status (elevated versus normal) using systolic and diastolic blood pressure levels at baseline. In accordance with American College of Cardiology/American Heart Association (ACC/AHA) guidelines, elevated blood pressure was defined as a systolic blood pressure level of at least 130 mmHg and/or a diastolic blood pressure level of at least 80 mmHg (Flack & Adekola, 2020). Only participants with both systolic and diastolic blood pressure levels below the threshold were assigned to the normal blood pressure group.

#### Cognitive Measures

Cognitive functioning across three domains was assessed using multiple neuropsychological tests. The three domains were memory, reasoning, and processing speed. Memory was assessed using the Hopkins Verbal Learning Test (HVLT), Rey Auditory-Verbal Learning Test (RAVLT), and Rivermead Behavioral Memory Test (RBMT) (Barbara et al., 1985; Brandt, 1991; Rey, 1941). The HVLT evaluates verbal learning and memory through immediate recall, delayed recall, and recognition of a 12-item word list. The RAVLT measures verbal learning and memory by assessing immediate and delayed recall of 15 unrelated words presented across multiple learning trials. The RBMT assesses everyday memory functioning through ecologically valid tasks designed to detect memory problems in daily life.

Reasoning was assessed with three tests: Word Series, Letter Series, and Letter Sets (Ekstrom & Harman, 1976; Gonda & Schaie, 1985; McArdle & Prindle, 2008; Thurstone & Thurstone, 1949; Willis & Caskie, 2013). The Word Series test measures inductive reasoning by requiring participants to identify patterns in word sequences and apply the rule to complete new sequences. The Letter Series evaluates logical thinking and pattern recognition through the identification of patterns in sequences of letters. Letter Sets assess inductive reasoning by requiring participants to identify which letter set follows a different pattern than others.

Processing speed was assessed with Digit Symbols Substitution (DSS), Digit Symbols Copy (DSC), and Useful Field of View (UFOV) (Ball et al., 1993; Wechsler, 1955). DSS measures psychomotor processing speed by requiring rapid matching of symbols to digits according to a provided key. DSC assesses pure motor speed by having participants copy symbols as quickly as possible without the matching component. UFOV evaluates the visual field over which information can be extracted at a brief glance, measuring visual attention and processing speed.

The cognitive tests were administered at each time point. The global cognition composite for each time point was calculated as the mean of the T-scores for each participant’s performance on all the cognitive tests at that time point. Domain-specific composites were also computed with the same approach using T-scores from corresponding cognitive tests. The memory composite comprised scores from HVLT, RAVLT, and RBMT. The composite for reasoning included scores from word series, letter series, and letter sets, while the processing speed composite included DSS, DSC, and UFOV. The raw scores for UFOV were reversed such that higher scores indicated better performance to align with the direction of the other cognitive measures.

### Statistical Analysis

Linear mixed-effects models were employed using SAS PROC MIXED with full maximum likelihood estimation to investigate the effect of pulse pressure on cognitive decline (Singer & Willett, 2003). Likelihood ratio tests were used to determine if adjusting for attrition and adding both fixed and random quadratic effects of time significantly improved the model fit. These adjustments were retained as they significantly improved model fit, better accounting for participant drop-out and non-linear changes over time (supplementary material).

#### The effect of pulse pressure on cognitive decline

To examine the effect of pulse pressure on the rate of decline in cognition, global cognition was regressed on time-varying pulse pressure, time (linear and quadratic), the interaction between pulse pressure and linear time, and covariates (GC = β0 + βpp + βtimeL + βtimeQ + βpp * timeL + βcovariates). The random effect of time (linear and quadratic) was also added to this model, as well as all other linear mixed-effect models in this study. The model was replicated for memory, reasoning, and processing speed to examine the effect of pulse pressure on the decline of these cognitive domains.

Covariates included age, education, race, sex, cognitive training status, and attrition. This set of covariates was also used in all subsequent models. Cognitive training status was a binary variable with participants who received cognitive training after the baseline assessment coded as one and those who did not receive any training coded as zero. Attrition was also indicated with a binary variable where participants who dropped out at any point in time were coded as one, and those who participated in all the time points were coded as zero.

#### The additional predictive benefit of pulse pressure over blood pressure status

To examine whether pulse pressure provides an additional predictive benefit of cognitive decline over blood pressure status, several models were evaluated. First, the previous model was adjusted to control for the effect of blood pressure status (GC = β0 + βpp + βbps + βtimeL + βtimeQ + βpp * timeL + βcovariates). This is to examine whether the effect of pulse pressure remains consistent after controlling for the effect of blood pressure status. To evaluate the collinearity of pulse pressure and blood pressure status, the variance inflation factor of the two variables was examined by including pulse pressure in an ordinary least squares regression model evaluating cognition at baseline (GC = β0 + βpp + βbps + βcovariates).

Second, a linear mixed-effects model with an interaction between blood pressure status and linear time (GC = β0 + βbps + βtimeL + βtimeQ+ βbps * timeL + βcovariates) examined whether blood pressure status is associated with cognitive decline. This was used to compare the predictive capacity of blood pressure status to that of pulse pressure on cognitive decline.

#### The differential effect of pulse pressure based on blood pressure status

To examine the differential effect of pulse pressure on cognitive decline based on blood pressure status, several models were evaluated. This included a linear mixed-effects model that included pulse pressure, blood pressure status, time, and all possible interactions among these variables (GC = β0 + βpp + βbps + βtimeL + βtimeQ + βpp * βtimeL + βbps * βtimeL + βpp * βbps + βpp *βtimeL* βbps + βcovariates). The emphasis was on the three-way interaction between pulse pressure, blood pressure status, and linear time (βpp *βtime* βbps), which indicated whether the effect of pulse pressure on cognitive decline varies based on blood pressure status.

The three-way interaction model was followed by a stratified model (GC = β0 + βpp + βtimeL + βtimeQ+ βpp * βtimeL + βcovariates) for the two blood pressure groups (normal versus elevated). This stratified model was replicated for the three cognitive domains to examine whether the negative effect of pulse pressure on these domains differs between the elevated and normal blood pressure status.

### Sensitivity Analyses

To ensure our findings are not limited to the chosen criterion for defining blood pressure status based on baseline blood pressure levels, the analysis was replicated with a different criterion. This other criterion averages blood pressure levels from all the time points to define blood pressure status. Additionally, blood pressure status in the models for the second aim was replaced with time-varying mean blood pressure, systolic blood pressure, and diastolic blood pressure in separate models to compare the predictive capacities of each of these measures against pulse pressure.

## Results

### Demographic Statistics

Descriptive statistics for all variables at baseline and whether these variables significantly differed between the normal and elevated blood pressure status are presented in Table 1. Older adults in the normal blood pressure group were more likely to be white, had more years of education, lower pulse pressure, and better cognitive status in global cognition, reasoning, and processing speed. All other variables did not significantly differ between the two groups.

**Table 1.** Comparison of Variables by Blood Pressure Status at Baseline (N = 2787)

| **Variable** | **Blood Pressure Status** |  | ***t* or $\chi^2$** |
| --- | --- | --- | --- |
|  | **Normal**<br>**(n = 979)** | **Elevated**<br>**(n=1808)** |  |
| Age (years): M (SD) | 73.67 (5.87) | 73.64 (5.94) | .13 |
| Education (years): M (SD) | 13.96 (2.68) | 13.30 (2.69) | 6.17** |
| Race: |  |  |  |
| White | 78.92% | 70.27% |  |
| Non-white | 21.08% | 29.73% | 24.19** |
| Sex: |  |  |  |
| Male | 22.47% | 25.06% |  |
| Female | 77.53% | 74.94% | 2.31 |
| Cognitive Training Status: |  |  |  |
| Cognitively Trained | 75.08 | 75.00 |  |
| Not Cognitively Trained | 24.92 | 25.00 | .00 |
| Pulse Pressure, M (SD) | 48.51(8.65) | 66.89 (15.94) | -39.00** |
| Global Cognition, M (SD) | 50.63 (7.05) | 49.67 (6.54) | 3.54** |
| Memory, M (SD) | 50.42 (8.67) | 49.80 (7.96) | 1.86 |
| Reasoning, M (SD) | 51.02 (9.28) | 49.47 (8.74) | 4.32** |
| Processing Speed, M (SD) | 50.45 (7.11) | 49.75 (6.75) | 2.56* |

*Note.* \* $p < .05$ ; \*\* $p < .001$

### The effect of pulse pressure on cognitive decline

Pulse pressure had a significant negative effect on the rate of decline in global cognition (*b* = −0.003, SE = 0.001, *t* = −3.92, *p* < 0.001) after accounting for age, education, race, sex, and cognitive training status. This suggests that higher pulse pressure is associated with a faster rate of decline in global cognition (Figure 1A).

**Figure 1.**
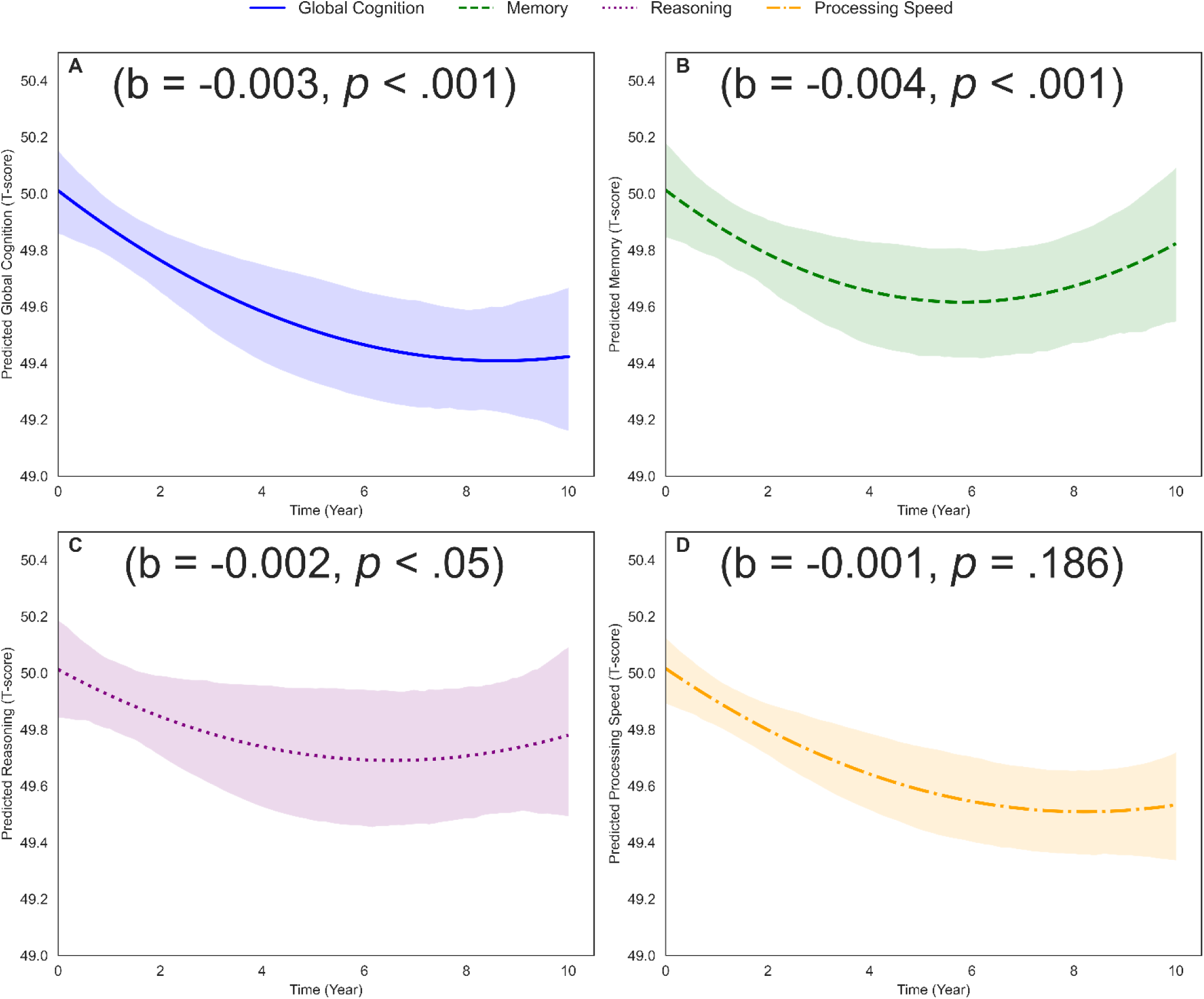
The effect of pulse pressure on global cognitive decline (A) and domain-specific cognitive decline (B - D).

Pulse pressure was also associated with the decline in specific cognitive domains (Figure 1B, 1C, and 1D). Specifically, pulse pressure was associated with an accelerated rate of decline in memory (*b* = −0.004, SE = 0.001, *t* = −3.66, *p* < .001) and reasoning (*b* = −0.002, SE = 0.001, *t* = −2.54, *p* < .05) after accounting for all covariates. However, the effect of pulse pressure on the decline in processing speed was non-significant (*b* = −0.001, SE = 0.001, *t* = −1.32, *p* = .186).

### The additional predictive benefit of pulse pressure over blood pressure status

The effect of pulse pressure on global cognitive decline was still observed as statistically significant, with a similar effect estimate after adjusting for the effect of blood pressure status (*b* = −0.003, SE = 0.001, *t* = −3.89, *p* < .001). This suggests that the effect of pulse pressure on cognitive decline is not confounded by blood pressure status. Adjusting for blood pressure status did not change the slope of the relationship between pulse pressure and cognition (Figure 2).

**Figure 2.**
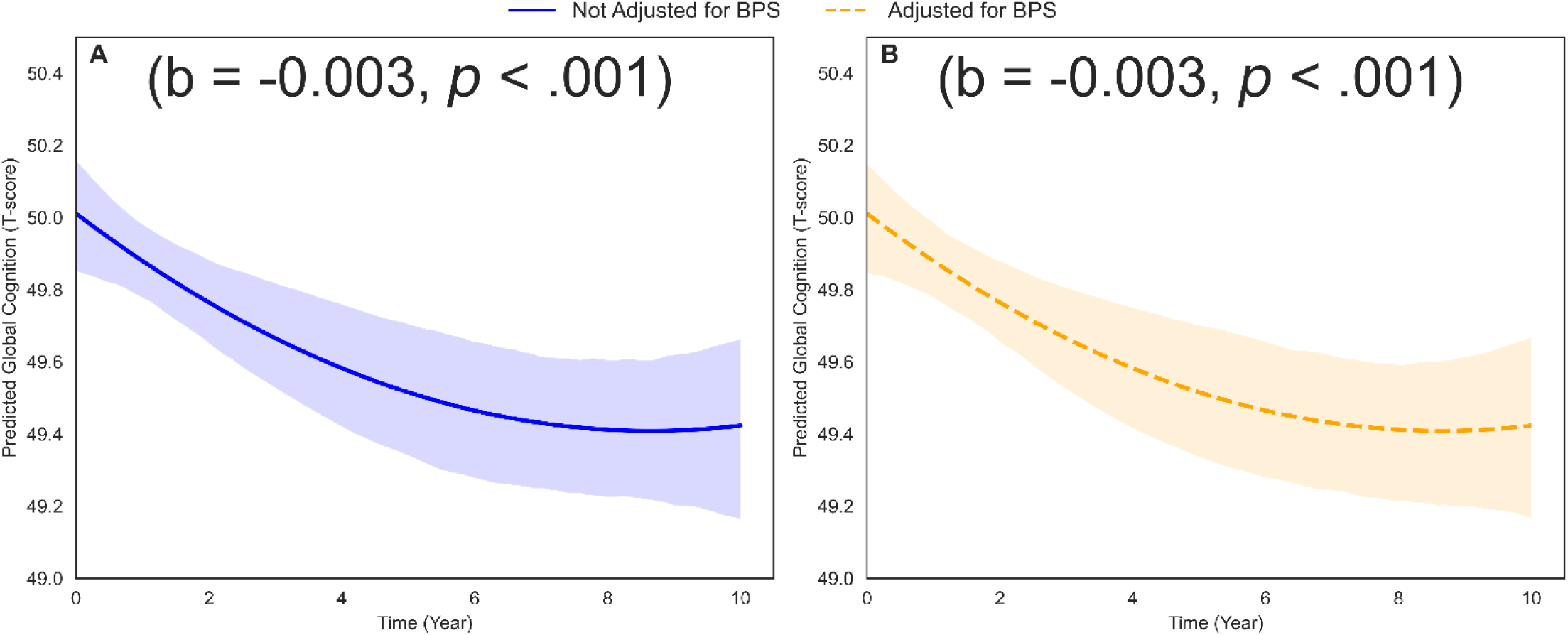
The effect of pulse pressure on global cognitive decline before (A) and after (B) adjusting for blood pressure status.

These effects of pulse pressure and blood pressure status were confirmed to be uncorrelated by the variance inflation factors of 1.20 and 1.17 for pulse pressure and blood pressure status, respectively. Sensitivity analyses also revealed a similar pattern, where controlling for time-varying mean blood pressure, systolic blood pressure, and diastolic blood pressure in separate models did not change the effect of pulse pressure on global cognitive decline (supplemental tables 1 through 3). The findings were also consistent when blood pressure status was defined by blood pressure levels at each time point (supplemental table 4).

Unlike pulse pressure, blood pressure status was not significantly associated with the rate of global cognitive decline (*b* = 0.019, SE = 0.028, *t* = 0.68, *p =* .495). This result was consistent across all three cognitive domains; blood pressure status was not significantly associated with the rate of decline in memory, reasoning, or processing speed (supplemental tables 5 through 7). This suggests that, in addition to the independent effects of pulse pressure and blood pressure status, pulse pressure provides an additional predictive utility beyond blood pressure status. This result was also consistent with the sensitivity analyses, where time-varying mean blood pressure, diastolic blood pressure, and blood pressure status based on average level across timepoints did not significantly predict cognitive decline (supplemental tables 8 through 10). Although systolic blood pressure significantly predicted global cognitive decline (*b* = −0.001, SE = 0.001, *t* = −2.34, *p* < .05) (supplemental table 11), the effect size was smaller relative to pulse pressure.

### The differential effect of pulse pressure based on blood pressure status

The three-way interaction between pulse pressure, time, and blood pressure status on global cognition was marginal (*b* = −0.003, SE = 0.002, *t* = −1.97, *p* = .050). This suggests that the effect of pulse pressure on cognitive decline may differ between the two blood pressure statuses (Figure 3).

**Figure 3.**
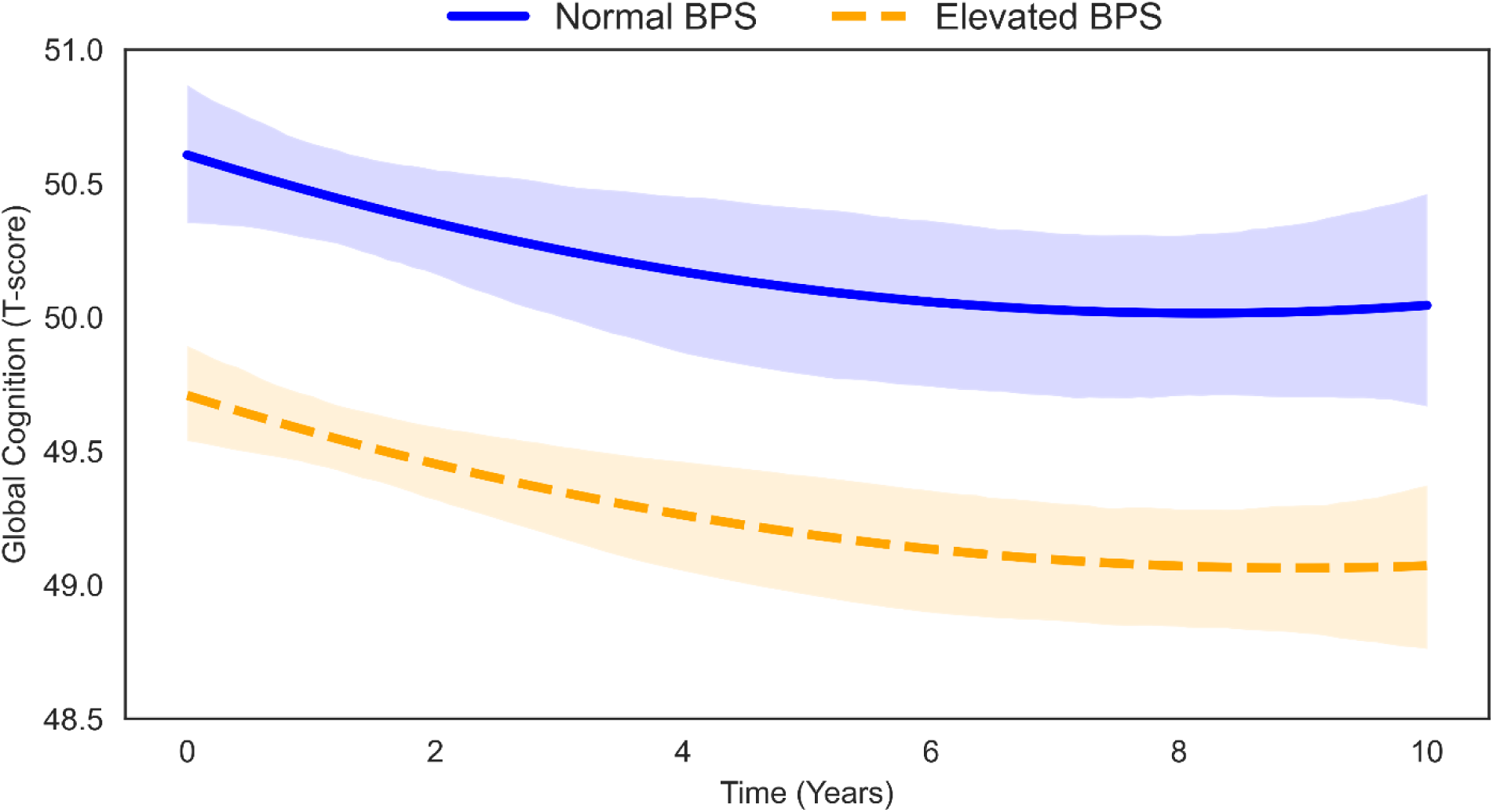
The effect of pulse pressure on global cognitive decline based on blood pressure status.

Stratification analyses confirmed the differential effect of pulse pressure on cognitive decline between the normal and elevated blood pressure groups. Specifically, the negative effect of pulse pressure on global cognitive decline was only significant in the elevated blood pressure group (*b* = −0.004, SE = 0.001, *t* = −4.56, *p* < .001), but not in the normal blood pressure group (*b* = −0.000, SE = 0.002, *t* = −0.29, *p* = .769). This pattern was consistent across all cognitive domains (Table 2). The results were generally consistent with those from sensitivity analyses, which used blood pressure status based on the average level across time points. Specifically, the differential effect was observed for global cognition, memory, and reasoning, but not for processing speed (supplemental tables 20 through 27). These results suggest that the acceleration of cognitive decline due to pulse pressure is only present among older adults with elevated blood pressure. This differential effect of pulse pressure on cognition is more apparent using quartiles of pulse pressure derived from the combined sample. Among the elevated blood pressure group, a predicted decline is evident across all the quartiles of pulse pressure. However, a more stable trajectory is observed for the normal blood pressure group, especially in the three lowest quartiles (Figure 4). This pattern was consistent across all specific cognitive domains (supplemental figures 1 through 3).

**Figure 4.**
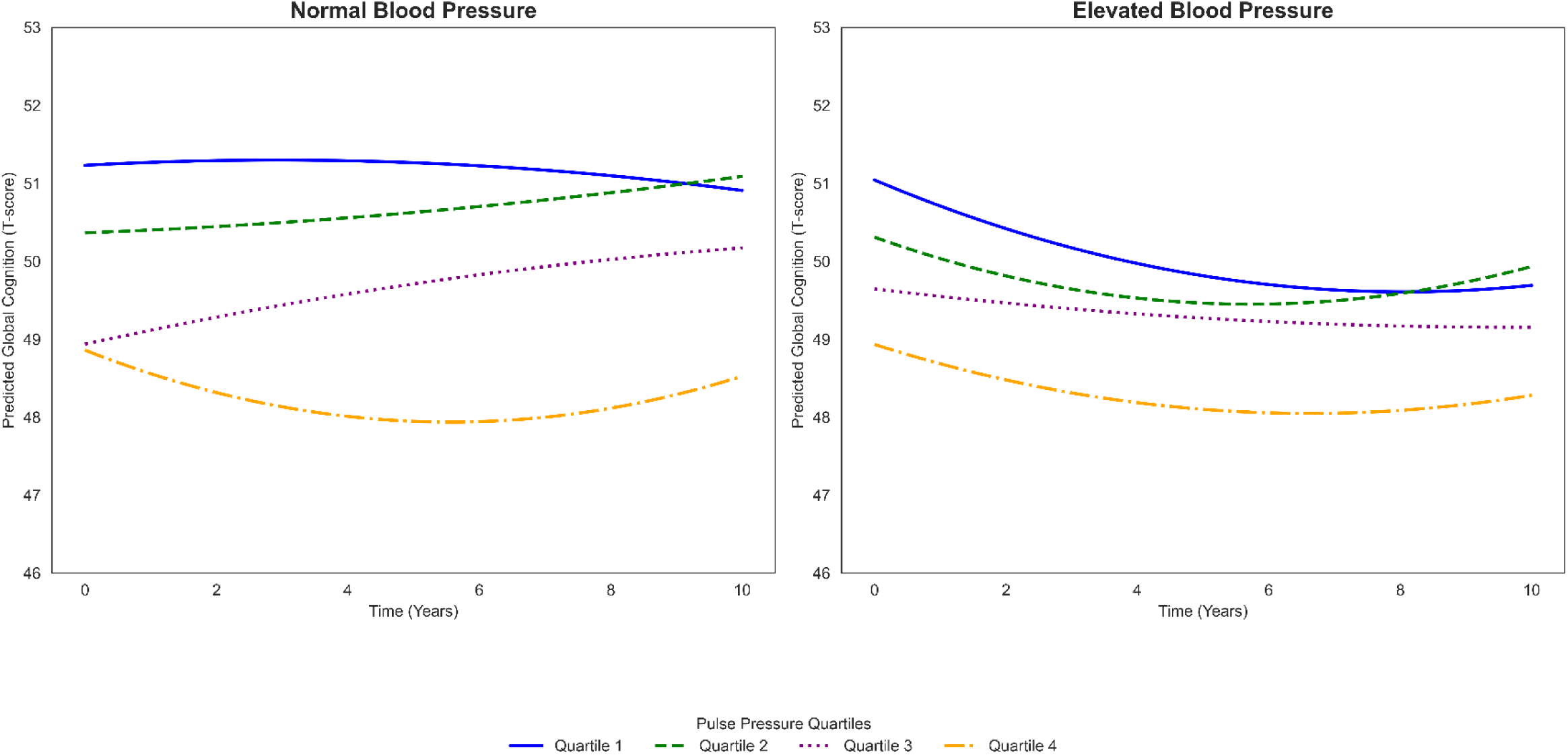
The effect of pulse pressure on global cognitive decline for normal (A) and elevated (B) blood pressure status. The separate lines within each blood pressure status represent the quartiles for pulse pressure, with 1 being the lowest and 4 being the highest quartile.

**Table 2.** Differential Effect of Pulse Pressure on Cognitive Decline Based on Blood Pressure.

| Effect (Slope) | Normal Blood Pressure |  |  |  | Elevated Blood Pressure |  |  |  |
| --- | --- | --- | --- | --- | --- | --- | --- | --- |
|  | Estimate | SE | *t* | *p* | Estimate | SE | *t* | *p* |
| Global Cognition | -0.000 | 0.002 | -0.29 | **0.769** | -0.004 | 0.001 | -4.56 | < **0.001** |
| Memory | -0.003 | 0.002 | -1.05 | **0.293** | -0.004 | 0.001 | -3.30 | **0.001** |
| Reasoning | -0.000 | 0.002 | -0.12 | **0.904** | -0.003 | 0.001 | -2.88 | **0.004** |
| Processing Speed | 0.000 | 0.002 | 0.19 | **0.849** | -0.003 | 0.001 | -2.23 | **0.027** |

*Note.* Refer to supplemental tables 12 through 19 for the complete output for each of the models in table 2

## Discussion

We investigated the longitudinal effect of pulse pressure on cognition among older adults, with a particular focus on how this relationship might be influenced by blood pressure status. The findings indicate that higher pulse pressure was significantly associated with a faster rate of decline in cognition, especially in memory and reasoning. The effect of pulse pressure on cognitive decline was not confounded by blood pressure status, and pulse pressure was a superior predictor of cognitive decline relative to blood pressure status. Additionally, the findings indicate a differential effect of pulse pressure based on blood pressure status, where the accelerated effect of pulse pressure on cognitive decline was only present among older adults with elevated blood pressure.

The observed association between higher pulse pressure and faster cognitive decline aligns with previous studies linking pulse pressure to cognitive aging (Nation et al., 2015; Shi et al., 2020; Zhou et al., 2023). Some neurobiological mechanisms can explain this association. Pulse pressure is a marker of arterial stiffness, which is associated with cerebrovascular damage through impaired autoregulation of cerebral perfusion (Thorin-Trescases et al., 2018). Cerebrovascular damage, including small vessel disease, directly affects cognition as well as promotes neurodegeneration to accelerate cognitive decline (Erdener & Dalkara, 2019; Rensma et al., 2020). These findings are consistent with studies linking higher pulse pressure to lower total cerebral volume (Nation et al., 2015), accelerated entorhinal and hippocampal atrophy, reduced white matter integrity (King et al., 2025), and higher phosphorylated tau and amyloid-beta burden (Shi et al., 2020).

The unique influence of high pulse pressure over elevated blood pressure on cognitive decline can also be understood through the autoregulation of cerebral perfusion. In hypertensive patients, autoregulation is adjusted by shifting the lower and upper thresholds to accommodate the chronically elevated levels of blood pressure (Strandgaard & Paulson, 1995). However, high pulse pressure can disrupt autoregulation without accommodation, intensifying the damage to microvasculature (Thorin-Trescases et al., 2018). This suggests the cerebrovascular system, through autoregulation, can accommodate the negative effect of elevated blood pressure more than that of high pulse pressure. Importantly, this hypertensive adaptation of autoregulation of cerebral perfusion makes the cerebrovascular system more vulnerable to injuries (Barry, 1985).

The observed differential effect of pulse pressure on cognitive decline based on blood pressure status further supports the increased susceptibility of the cerebrovascular system due to hypertensive adaptation of cerebral perfusion autoregulation. While this hypertensive adaptation protects the brain from high pressures, it increases the brain’s susceptibility to ischemic damage at low pressures (Barry, 1985). Hypertension is associated with the incidence and progression of cerebral small vessel disease, with marked white matter hyperintensities and cerebral microbleeds (Liu et al., 2018; Nam et al., 2019; Petrea et al., 2020). These potentially account for the increased susceptibility only among older adults, specifically with elevated blood pressure, facilitating the observed acceleration by high pulse pressure on cognitive decline. This aligns with the findings of Riba-Llena et al. (2016), where high pulse pressure was associated with the presence of deep white matter hyperintensities and mild cognitive impairment among hypertensive older adults. Perhaps without elevated blood pressure, the cerebral vascular system better accommodates the effects of increased pulse pressure, thereby mitigating its negative effect on cognition. This work provides further evidence that health and medical illness, independent of dementia, are related to cognitive decline (Schretlen et al., 2024), and that hypertension treatment and pulse pressure monitoring can help reduce the risk of cognitive decline and dementia (Walker et al., 2017).

Some limitations should be considered in interpreting the findings of this study. First, while the associations between pulse pressure and cognitive decline were statistically significant, the observed effect sizes were small after adjusting for several relevant covariates. This suggests pulse pressure is one of many factors contributing to cognitive decline among older adults. Second, blood pressure status was based on blood pressure assessment, not a hypertension diagnosis. We chose this approach because some participants with a diagnosis of hypertension could be well-treated and have normal blood pressure, and some participants could have elevated blood pressure that was undiagnosed. Thus, using blood pressure assessments provides the opportunity to accurately categorize participants based on their true state, not influenced by misdiagnosis or hypertensive medications. Finally, blood pressure measurements were taken at discrete time points and may not fully capture daily variations in cardiovascular function. Future studies could utilize ambulatory blood pressure monitoring for more comprehensive data.

### Perspectives

The differential impact of pulse pressure on cognitive decline based on blood pressure status revealed in this study has significant implications for clinical practice and future research. These findings demonstrate that pulse pressure could serve as a valuable biomarker when determining cognitive risk, particularly among older adults with elevated blood pressure. Incorporating pulse pressure monitoring into routine geriatric assessments may enhance early identification of individuals at heightened risk for cognitive decline, potentially enabling timely interventions. Future research should investigate the neurobiological mechanisms underlying the interaction between pulse pressure and elevated blood pressure on cerebrovascular function to help advance our understanding of vascular contributions to cognitive impairment. Additionally, examining whether this differential effect extends to clinical populations with mild cognitive impairment or early dementia would further clarify the clinical utility of pulse pressure as a prognostic marker. The findings highlight the need to account for blood pressure status in studies investigating the effect of pulse pressure on cognitive function.

### Conclusion

The findings demonstrate that higher pulse pressure is significantly associated with accelerated cognitive decline, and pulse pressure offers a complementary predictive utility of blood pressure status on cognitive decline. Notably, the accelerated cognitive decline associated with increased pulse pressure was significant only among older adults with elevated blood pressure, suggesting a synergistic relationship between arterial stiffness and hypertension, which may exacerbate cerebrovascular damage. These findings have important clinical implications, as monitoring pulse pressure, particularly in individuals with elevated blood pressure, could enhance identification of those at heightened risk for cognitive decline. The findings also underscore the importance of blood pressure management among older adults to mitigate the effect of pulse pressure on cognition.

